# Increased circulating levels of angiotensin-(1-7) in severely ill COVID-19 patients

**DOI:** 10.1101/2021.02.01.20232785

**Authors:** Ana Luiza Valle Martins, Filipe Alex da Silva, Lucas Bolais-Ramos, Gisele Capanema de Oliveira, Renata Cunha Ribeiro, Danilo Augusto Alves Pereira, Filippo Annoni, Mirella Monique Lana Diniz, Thuanny Granato Fonseca Silva, Bruna Zivianni, Alexandre Carvalho Cardoso, Juliana Carvalho Martins, Daisy Motta-Santos, Maria José Campagnole-Santos, Fabio Silvio Taccone, Thiago Verano-Braga, Robson Augusto Souza Santos

## Abstract

The mono-carboxypeptidase Angiotensin-Converting Enzyme 2 (ACE2) is an important “player” of the renin-angiotensin system (RAS). ACE2 is also the receptor for SARS-CoV-2, the new coronavirus that causes COVID-19. It has been hypothesized that following SARS-CoV-2/ACE2 internalization Ang II level would increase in parallel to a decrease of Ang-(1-7) in COVID-19 patients. In this preliminary report, we analyzed the plasma levels of angiotensin peptides in 19 severe COVID-19 patients and 19 non-COVID-19 volunteers, to assess potential outcome associations. Unexpectedly, a significant increase in circulating Ang-(1-7) and lower Ang II plasma level were found in critically ill COVID-19 patients. Accordingly, an increased Ang-(1-7)/ Ang II ratio was observed in COVID-19 suggesting a RAS dysregulation toward an increased formation of Ang-(1-7) in these patients.

## Introduction

The mono-carboxypeptidase Angiotensin-Converting Enzyme 2 (ACE2) is a major player in the Renin-Angiotensin System (RAS) as it converts the decapeptide Angiotensin I (Ang I) to Ang-(1-9), and Angiotensin II (Ang II) to Ang-(1-7) (Figure 1A).^1^ ACE2 is also a target for the new human coronavirus SARS-CoV-2, which is responsible for the dramatic COVID-19 ongoing pandemic.^2^ It has been suggested that following SARS-CoV-2/ACE2 internalization, Ang II level increases^3^ in parallel to a decrease of Ang-(1-7) level.^4^ These changes would be expected both at tissue and circulatory levels. Considering that Ang-(1-7) has many beneficial effects, including anti-inflammatory, anti-thrombogenic and anti-fibrotic activities,^1^ it has been hypothesized that Ang-(1-7) administration would improve the clinical outcome of COVID-19 patients. Aiming to test this hypothesis, a Phase I/II clinical trials (NCT04633772) has been initiated with a planned Phase III clinical trial (NCT04332666).

**Figure 1.**
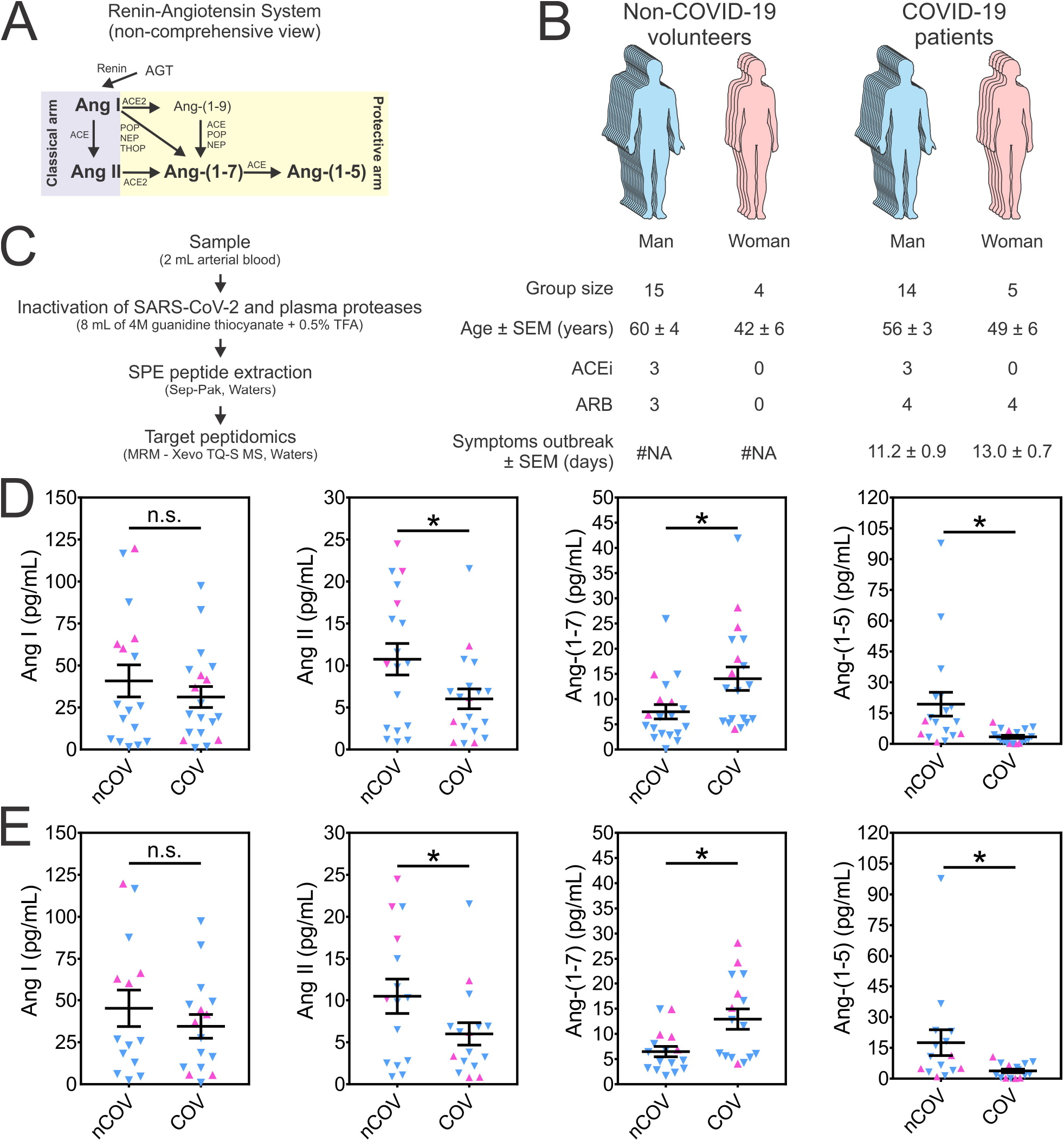
Circulating renin-angiotensin peptides in SARS-CoV-2 infection. (**A**) A non-comprehensive view of the formation of the peptides from the RAS. Peptides measured in this study are in bold. (**B**) Epidemiologic parameters of the subjects included in this study (COVID-19 patients and non-COVID-19 volunteers). (**C**) A simplified schematic view of the methodology employed to quantify the selected peptides from the RAS. (**D**) Arterial blood concentration of the selected RAS peptides (Ang I, Ang II, Ang-(1-7) and Ang-(1-5)) from COVID-19 patients and non-COVID-19 volunteers. (**E**) Arterial blood concentration of the RAS peptides excluding individuals under ACEi treatment. Man is represented by (▼) and woman by (▲). Data are presented as mean ± SEM. Parametric t test was used for the statistical analyses. ^*^*P* < 0.05. Abbreviations: ACE, angiotensin-converting enzyme; ACE2, angiotensin-converting enzyme 2; ACEi, ACE inhibitor; AGT, angiotensinogen; ARB, Ang II receptor (AT1) antagonist; COV, COVID-19 patients; MRM, multiple reaction monitoring; nCOV, non-COVID-19 volunteers; NEP, neutral endopeptidase; POP, prolyl oligopeptidase; RAS, reninangiotensin system; SPE, solid phase extraction; THOP, thimet oligopeptidase.

This report aims at answering the key question whether the circulating levels of angiotensin peptides are indeed altered in COVID-19 patients using LC/MS/MS (Figure 1B). This is important since that there is no data in the literature on the concentration of circulating angiotensin using direct measurement of these peptides in COVID-19 patients. Others reported that Ang II is higher in the plasma of COVID-19 patients, but the authors used an ELISA method to quantify Ang II.,^3^ This method has been recently criticized by Chappell et al. due to its poor specificity for the measurement of Ang II and Ang-(1-7) in human plasma.^5^ Also, Kintscher et al. have recently reported the “plasma angiotensin peptide profiling” in COVID-19 patients^6^ but, even though they used a mass spectrometry-based strategy, they collect the blood using neither protease inhibitors nor under denaturation conditions to “quench” the activity of plasma proteases. Actually, they even incubated the heparinized plasma samples at 37° C to increase proteases activities and thus measure what they call “peptides’ equilibrium concentrations”. This method allows only the indirect measurement of RAS peptides as it does not account for the important role of enzymes on the endothelium, for example, in production or hydrolysis of RAS peptides in the circulation. Here, we are reporting the direct measurements of the RAS peptides Ang I, Ang II, Ang-(1-7) and Ang-(1-5) in the arterial blood from 19 COVID-19 patients and 19 non-COVID-19 volunteers.

## Methods

The study protocol has been approved by the Ethics Committee of the Federal University of Minas Gerais, Belo Horizonte, Brazil (CAAE 34080720.0.1001.5149). Written consents were obtained from the patients or their relatives for blood sampling. The COVID-19 patients were recruited in two hospitals (Mater Dei Hospital and Eduardo de Menezes Hospital, Belo Horizonte, Minas Gerais, Brazil). Arterial blood samples were obtained from patients admitted at the hospitals’ Intensive Care Unit (ICU) upon admission and before starting the any additional treatment (medications, oxygen, etc.). The arterial samples from non-COVID-19 subjects were collected from healthy volunteers (n=6) or before cardiac catheterization (n=13). Importantly, only the arterial blood samples from patients with no diagnostic for cardiovascular diseases, *as per* the catheterization results, were used in the study. Three patients and three non-COVID-19 individuals were previously using ACE inhibitors (ACEi) (Figure 1B) and the treatment was not interrupted at any time.

A mass spectrometry-based approach was used to quantify the RAS peptides. Inactivation of plasma proteases and potential SARS-CoV-2 in the blood samples was achieved by mixing 2 mL of arterial blood sample with 8 mL of a denaturation solution (4M guanidine thiocyanate in 0.5 % TFA). Samples (1,200 µL) were cleaned up using C18 solid-phase extraction (Sep-Pak, Waters), reconstituted in 60 µL 0.1% formic acid (concentration factor = 20-fold), and analyzed by LC-MS/MS (Xevo TQ-S, Waters) using the multiple reaction monitoring (MRM) mode (Figure 1C). The calibration curve was obtained using a stock solution containing synthetic peptides (Bachem, CA) of all RAS peptides used in this study. The applied calibration curve model (y = ax + b) proved accurate over the concentration range 10 to 1000 pg/mL (r = 0.997). The limit of quantitation (LOQ), inter- and intra-variability of this method has been previously reported.^7^ Data are shown as mean ± SEM and the parametric t test was used for the statistical analyses.

## Results

Arterial concentrations of the angiotensin peptides are shown in Figure 1D. In COVID-19 patients, the arterial concentration of Ang II (6.03 ± 1.18 vs.10.7 ± 1.87 pg/mL; *P* = 0.0381) and Ang-(1-5) (3.43 ± 0.75 vs.19.3 ± 5.80 pg/mL; *P* = 0.0084) were significantly lower than in the non-COVID-19 volunteers. Surprisingly, the blood levels of Ang-(1-7) were significantly higher in COVID-19 patients (14.0 ± 2.32 vs. 7.49 ± 1.42 pg/mL; *P* = 0.0214). No significant difference was observed for Ang I (31.2 ± 6.23 pg/mL vs. 40.8 ± 9.54 pg/mL; *P* = 0.3959). ACEi therapy did not significantly change the observed results, as shown by the calculated values excluding the data from individuals under ACEi treatment (Figure 1E): Ang I = 34.5 ± 7.07 vs. 45.2 ± 10.8 pg/mL; *P* = 0.4023; Ang II = 6.00 ± 1.33 vs. 11.2 ± 2.07; *P* = 0.0407; Ang-(1-7) = 12.9 ± 2.02 vs. 6.47 ± 1.03 pg/mL; *P* = 0.0080; Ang-(1-5) = 3.75 ± 0.86 vs. 17.5 ± 6.29 pg/mL; *P* = 0.0330.

## Discussion

Although we did not measure the tissular levels of RAS peptides, our findings contrast with the initial hypothesis that the interaction of SARS-CoV-2 with ACE2 would result in higher Ang II and lower Ang-(1-7) levels compared to non-COVID-19 subjects.^4^ Indeed, a recent study using the equilibrium method^8^ is in line with our direct data. Previous studies suggested that one of the main routes to produce Ang-(1-7) in the circulation is ACE2-independent,^9, 10^ which may also explain our findings in the blood of COVID-19 patients. Significant decrease of Ang II and increase of Ang-(1-7) arterial levels were observed in critically ill COVID-19 patients (Figure 1D) and this is probably due to a direct dysregulation of RAS pathways in COVID-19 rather than a direct consequence of ACEi usage (Figure 1E). The Ang-(1-7)/Ang II ratio, which is an estimation of Ang II → Ang-(1-7) conversion, was 3-fold higher in cohort of COVID-19 patients (0.878 ± 0.201 vs. 2.79 ± 0.682; *P* = 0.0141), which may suggest an increased ACE2 or other Ang-(1-7)-forming activity in COVID-19. Indeed, an increase in soluble ACE2 in critically ill COVID-19 patients have been recently reported.^11^

In contrast to a previous report,^3^ we observed a significative reduction of Ang II concentration in COVID-19 patients, which may add more data against the reliability of ELISA to measure Ang II in human plasma.^5^ The Ang II/Ang I ratio was not significantly altered in the COVID-19 patients (0.292 ± 0.0517 vs. 0.205 ± 0.0322; *P* = 0.1518), which may suggest that ACE activity is potentially not altered in COVID-19. Unfortunately, we were unable to perform enzymatic activity assays in this study due to the denaturation conditions that we used to collect the samples.

Finally, with all the limitations of the small cohort included in this study, this is the first report of the direct measurement of RAS peptides in the circulation of COVID-19 patients. Although future studies are obviously necessary to better understand the effects of this disease on RAS pathways, our data provide new insights for the interpretation and planning of future therapies to modulate the RAS in the context of COVID-19.

## Data Availability

We willing to provide the raw data upon request

